# Role of hyper-reflective spots in predicting outcomes of intravitreal therapy in diabetic macular edema: A systematic review and meta-analysis

**DOI:** 10.1101/2021.04.16.21255622

**Authors:** Pratyusha Ganne, Nagesha C Krishnappa, Ganne Chaitanya, Siddharth K Karthikeyan

## Abstract

**Purpose:** Predicting response to intravitreal therapy in DME has become a challenging task. Individual studies have shown that HRS could be a reliable biomarker. This systematic review aimed to determine if there was a quantitative reduction in hyperreflective spots (HRS) following intravitreal therapy in diabetic macular edema (DME), if the type of intravitreal therapy (anti-VEGF versus steroid) had differential effects on quantitative HRS change and finally, if HRS at the start of therapy was associated with improvement in visual acuity (VA) or reduction in central macular thickness (CMT). We also aimed at bringing out the lacunae in the existing literature on HRS in DME and propose goals for future studies.

**Methods:** PubMed/MEDLINE, Scopus, ProQuest, CINAHL, Wiley online and Web of Science were searched based on MOOSE guidelines for non-randomized studies evaluating HRS as a biomarker in DME (between 1^st^ January 2011 and 1^st^ July 2020). Publication bias was analyzed using Begg and Mazumdar rank correlation test and funnel plots. Heterogeneity was assessed using the I^2^ statistic. Meta-analysis was done using a random-effects model.

**Results:** A total of 1168 eyes from 19 studies were eligible for inclusion. Pooled standardized mean differences showed that intravitreal therapy was associated with a reduction in quantitative HRS (z=-6.3, CI_95%=_-1.09 to −0.55, p<0.0001). Extreme between-study heterogeneity was observed (I^2=^93.2%) with significant publication bias. There was no difference in outcomes between anti-VEGF and steroid therapies (p=0.23). No definite conclusions could be drawn regarding the predictive value of HRS in determining the final VA and CMT.

**Conclusion:** This review could conclude that there is a definite reduction in quantitative HRS following either form of intravitreal therapy. Our conclusion about the role of HRS in predicting visual outcome and CMT change was limited by the number of analyzable studies owing to the wide variation in the study designs, methods and reporting.

## Introduction

Intravitreal anti-vascular endothelial growth factor (anti-VEGF) therapy has emerged as the first-line treatment for diabetic macular edema (DME) in the last decade after the landmark RISE/RIDE trials and Diabetic Retinopathy Clinical Research Network (DRCR.net) studies demonstrated a significant visual acuity (VA) improvement in as many as 60% of the eyes treated with these injections.^1,2^ However, as much as 50% of the eyes in protocols I and T of DRCR.net did not respond adequately to these injections.^3,4^ Intravitreal steroids in the form of dexamethasone implants are being used in such patients not responding to anti-VEGF injections.^5,6^ The rationale behind using steroids stems from the assumption that inflammation plays a significant role in some eyes with DME.^7,8^ However, a small subset of patients can show suboptimal response to dexamethasone implants as well.^9^ In a real-life scenario, predicting which patient will or will not respond to intravitreal treatment has become a challenging task. Hence, a lot of research is being done to understand what factors at baseline predict functional and morphological responses to these injections and which sub-groups of patients benefit from intravitreal steroids/ anti-VGEF injections.

A number of biomarkers are being evaluated on optical coherence tomography (OCT) scans to predict responses like neurosensory detachment (NSD),^10,11^ hyperreflective spots (HRS),^12^ ellipsoid zone (EZ) line integrity, cystoid macular edema (CME)^10^ and disorganization of retinal inner layers (DRIL).^13^ HRS are small, dot-like lesions that are of equal or higher reflectivity than the retinal pigment epithelium (RPE) / nerve fiber layer with absent back shadowing on OCT.^14-17^ Several theories have been postulated to explain the pathogenesis of these spots but their exact nature is still unclear. These spots are thought to be extravasated lipoproteins (precursors of hard exudates),^18^ inflammatory cells (leucocytes, activated microglia),^19,20^ migrated RPE cells^21^ or photoreceptor fragments.^22^ Significant research is underway to understand the predictive value of this biomarker in determining the final VA, reduction in central macular thickness (CMT), recurrence patterns and duration of action of intravitreal implants.^14,15,22-26^

The majority of the current literature on HRS in DME consists of small retrospective/prospective cohort studies. Though some studies have shown conflicting results, it seems promising that HRS could be used as a reliable marker of disease burden and response to therapy in DME. Hence, we tried to synthesize the available information on HRS to (1) investigate if there was a reduction in quantitative HRS following intravitreal therapy (2) determine if the type of intravitreal therapy (anti-VEGF vs. steroid) had differential effects on quantitative HRS change, and finally (3) if change in post-treatment quantitative HRS/ baseline HRS counts were associated with improvement in VA or reduction in CMT. Finally, we would like to draw attention to the lacunae in the existing literature on HRS in DME and suggest goals for future studies so that the true potential of this biomarker can be understood.

## Methods

We conducted this systematic review in accordance with Meta-Analyses and Systematic Reviews of Observational Studies (MOOSE) guidelines.^27^ Institutional review board exemption was obtained. The protocol has been registered in the International Prospective Register of Systematic Reviews (PROSPERO) (CRD42020186820).

### Selection criteria

This review included all articles that described HRS as an outcome predictor after intravitreal therapy in DME from peer reviewed journals published in the electronic databases (between 1^st^January 2011 and 1^st^ July 2020).

We excluded studies: (1) not available in English, (2) published in books or grey literature, conference abstracts, review, comments, letter to editor, case series (<5 subjects) (3) with insufficient quality (4) where the results of DME were combined with other causes of macular edema like vein occlusion (5) where additional interventions were done during the study period like laser, vitrectomy, etc. (6) performed in non-human subjects (7) where time domain OCT machines were used.

### Search Strategy

The following databases were searched: PubMed/MEDLINE, Scopus, ProQuest, CINAHL, Wiley online and Web of Science. PICO (participants, intervention, and comparison and outcomes) format search strategy was used to search the databases mentioned.

Key words used included: DMO, DME, diabetic retinopathy, DR, macular edema, macular odema, bevacizumab, anti-VEGF, ranibizumab, antivascular endothelial growth factor, BVZ, dexamethasone implant, steroid, intravitreal, intra-vitreal, injection, avastin, lucentis, accentrix, aflibercept, eyelea, ozurdex, triamcinolone acetonide, IVTA, steroid, conbercept, optical coherence tomography, SD-OCT, OCT, hyperreflective, hyper-reflective, hyper reflective, spots, foci, dots, material, points, aggregates, particles, clumps, elements, HRF, HS, HRS, HF, HRD, inflammatory biomarkers, prognostic markers, central macular thickness, CMT, CST, macular volume, foveal thickness, FT, best corrected visual acuity, VA, BCVA, outcomes. The full search strategy for MEDLINE using keywords is detailed in Appendix 1.

All titles and abstracts were pooled to a reference management database and duplicates removed. All titles and abstracts were independently screened by two board certified vitreo-retinal surgeons (PG and NCK) and full text copies of all potentially relevant papers were retrieved. Any conflict was resolved by mutual discussion. The reference list of each selected article was reviewed to ensure no relevant article was overlooked.

### Data extraction

Data from the final full-text articles was extracted by PG and was verified by NCK. Any conflicts were resolved by mutual discussion. The following data was extracted: (1) title, main author and publication year, (2) study design and sample size, (3) participant data, (4) intravitreal drug, (5) age, (6) gender distribution, (7) OCT machine used, (8) VA, (9) confounding factors (10) follow-up (11) area of the OCT scan over which HRS were analyzed, (12) HRS count before and after treatment, (13) association (i.e, correlation/ regression) between HRS, VA and CMT.

### Assessment of methodological quality and risk of bias

The quality and risk of bias of the articles included in full-text review was assessed by PG and SK using the National Institute of health (NIH) Study Quality Assessment Tool.^28^ Questions with answer ‘yes’ were scored as 1 and answer ‘no’ / ‘cannot determined’ / ‘not reported’ as 0. Any conflict was resolved by mutual discussion. Total score for each study = (the total number of questions with ‘yes’ as an answer / the total number of questions) x 100 (Note: If the answer to a question was ‘not applicable’, the question was excluded from both the numerator and the denominator). Studies were graded as high quality (80-100%), moderate quality (60-80%) and low quality (<60%).

### Statistical Analysis

#### Meta-analysis

For the purposes of completion of this systematic review, we performed a random-effects meta-analysis, whilst being aware of the need to explore possible heterogeneity of result-reporting in the various studies, as the topic is not thoroughly investigated as yet and also considering that fact that these studies may not all be randomized controlled trials or studies that bag Level-II evidence or higher. All the outcomes of interest (i.e. quantitative HRS reduction, difference in quantitative HRS reduction between steroid and anti-VEGF therapy, and post-treatment change in VA) were continuous variables. After integrating and synthesizing effect sizes from different studies, the variance of combined true effect sizes among the studies were estimated using Hedge’s g for all outcomes (with 95% CI). Random-effects model was used to account for heterogeneity between studies. Proportion of the variability that was due to heterogeneity was estimated using I^2^ statistics. Prediction interval (PI) of the combined effect size was estimated. Sub-group analysis was performed using ANOVA of sum of squares.^29^

Pre/post-treatment HRS counts (mean±SD) were parsed for meta-analysis to the test effect of intravitreal injections, following which a subgroup analysis was performed to differentiate outcomes of anti-VEGF and steroid sub-groups. Mean and SD of change scores of VA pre and post-treatment were parsed. We used imputation method to estimate SD of change scores when not explicitly reported by authors.

Publication bias was analyzed using Begg and Mazumdar rank correlation test (Δx-y, Kendall Tau a, PI limits and CI limits) and funnel plots.

## Results

### Included studies

Figure 1 shows the flow diagram to summarize inclusion of relevant studies.

**Figure 1:**
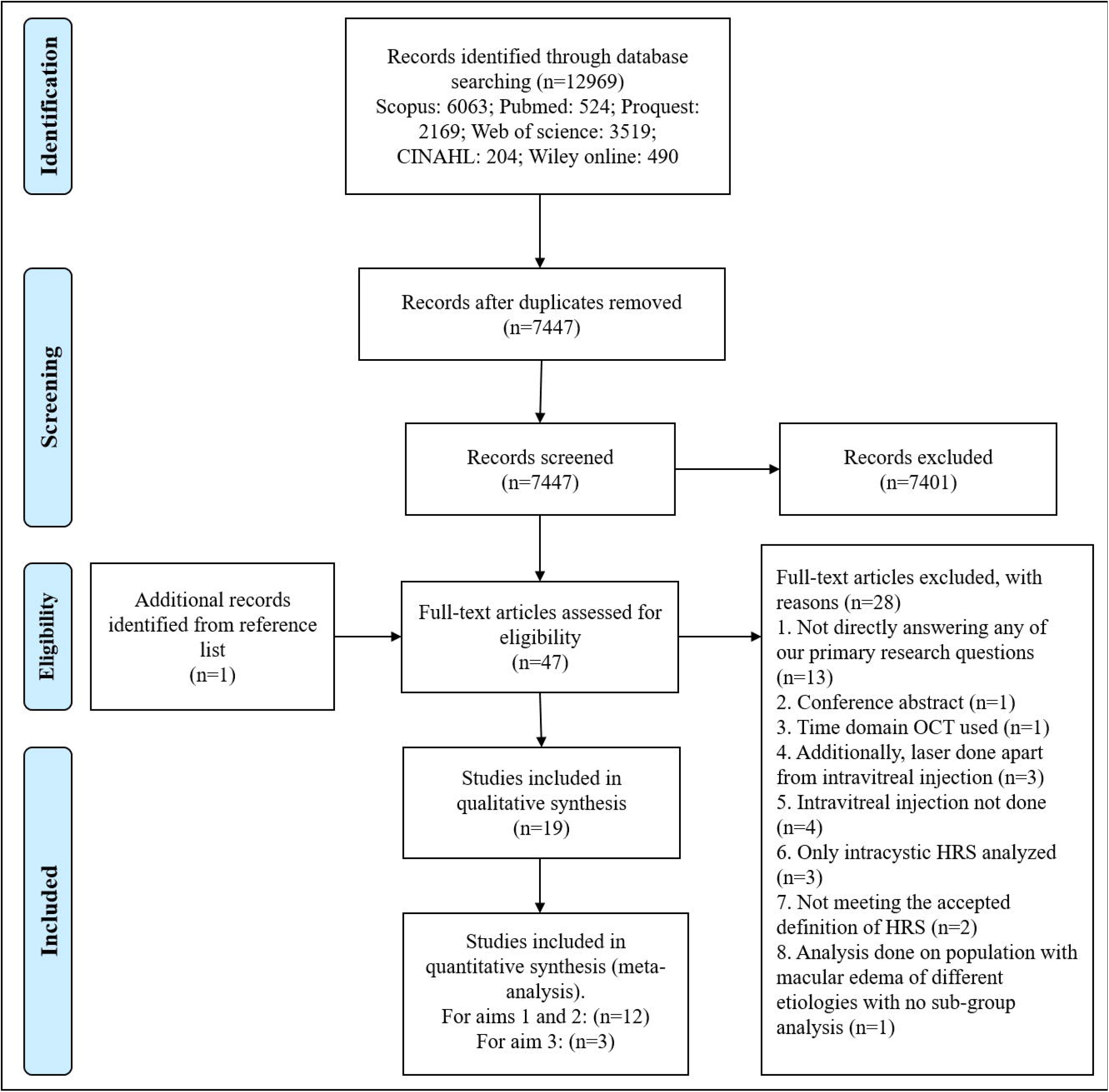
Flow diagram to summarize inclusion of relevant studies

### Quality of the studies

Nineteen studies were identified (thirteen were retrospective cohort studies,^12,15-17,24,26,30-36^ three were prospective cohort studies,^37,38,41^ two were case series^14,39^ and one was a case-control study^40^). The quality score of the studies is shown in Table 1.

**Table 1:**
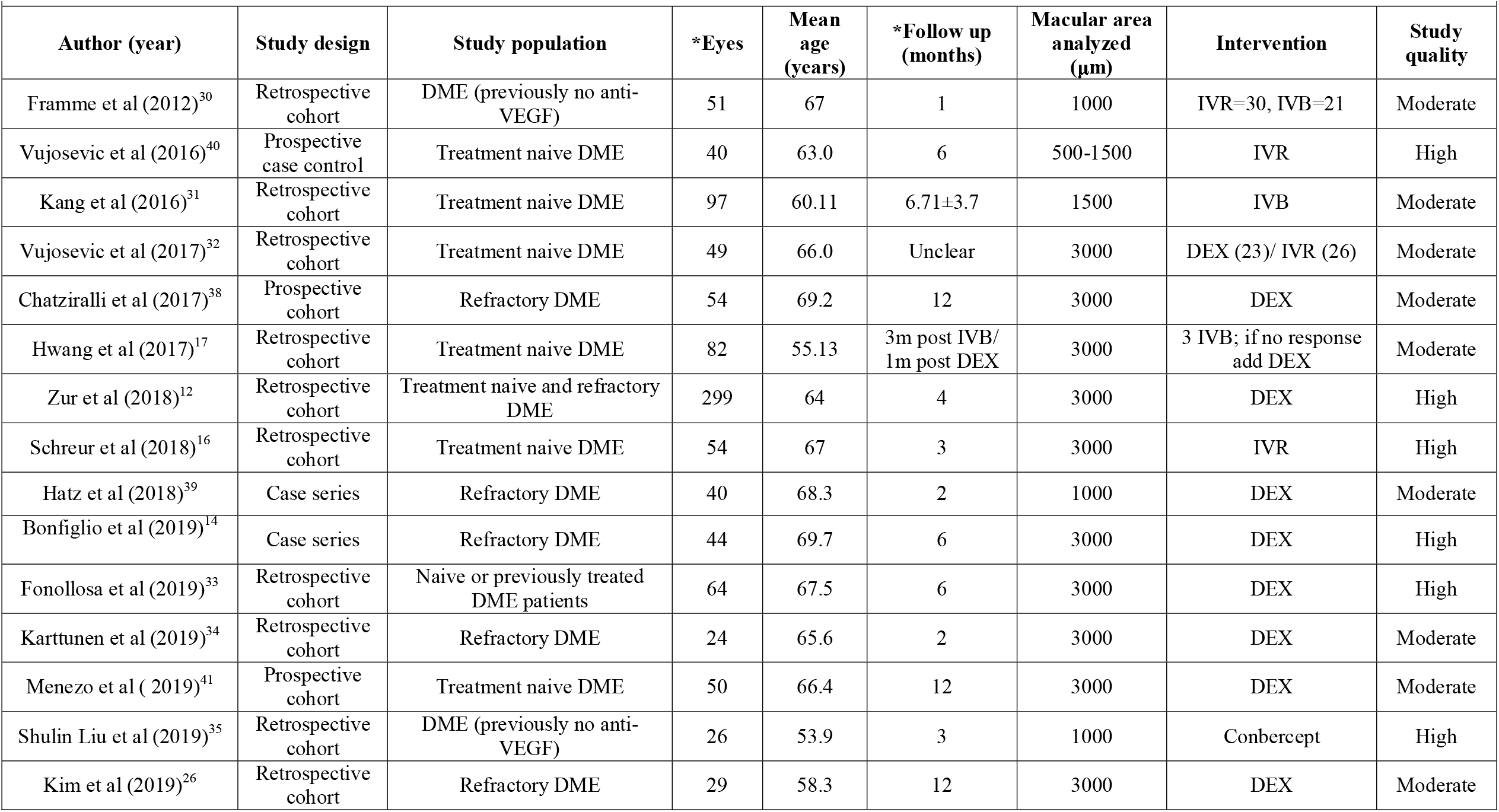
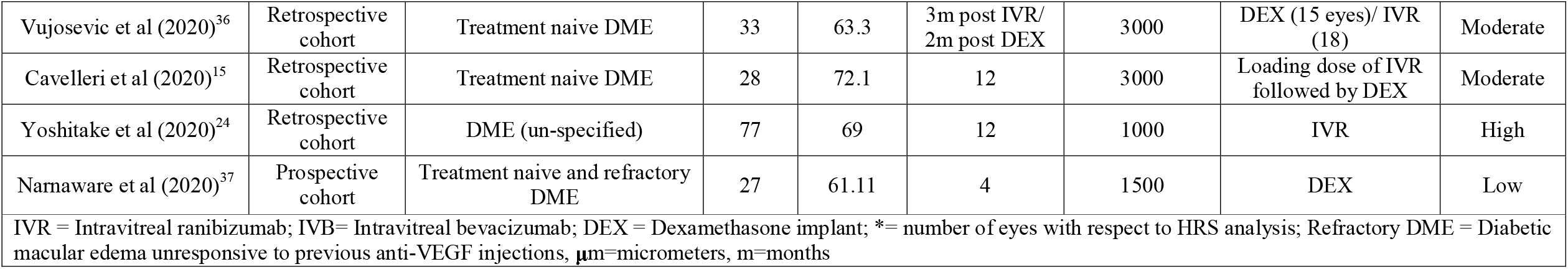
Baseline characteristics of the studies and participants included in the systematic review.

### Baseline characteristics (Table 1)

A total of 1168 eyes of 942 patients (mean age: 64.3±4.9 years, males: 59.4%) were analyzed for HRS from the above 19 studies. Eight studies evaluated the response to anti-VEGF injections [intravitreal ranibizumab (IVR), intravitreal bevacizumab (IVB), and Conbercept],^16,24,30-32,35,36,40^ 11 studies to dexamethasone implant^12,14,26,32-34,36-39,41^ and 2 studies to the sequential use of anti-VEGF and dexamethasone.^15,17^ The model of spectral domain OCT used, varied among the studies: 14 used SpectralisOCT™ (Heidelberg Engineering, Heidelberg, Germany),^12,14-17,24,26,30-32,34,35,38,39^ 3 used Cirrus HD-OCT™ (Carl Zeiss Meditec, Dublin, CA, USA),^33,37,41^ 1 each Retina scan™ (RS-3000 advance; Nidek)^40^ and DRI-SSOCT Triton plus™ (Topcon Medical Systems Europe, Milano, Italy).^36^ The measurement of HRS was done over different area sizes in the macula (12 studies used 3000µm area,^12,14-17,26,32-34,36,38,41^ 4 studies used 1000µm area,^24,30,35,39^ 2 studies used 1500µm area^31,37^ and 1 study used area between 500-1500µm from the center of the fovea).^40^

### Change in quantitative HRS with intravitreal therapy (Table 2)

Studies where HRS counts (mean±SD) were available before and after treatment were included in this analysis. There were a total of 12 studies. Quantitative HRS change with anti-VEGF and dexamethasone injections was reported in seven studies each.

**Table 2:**
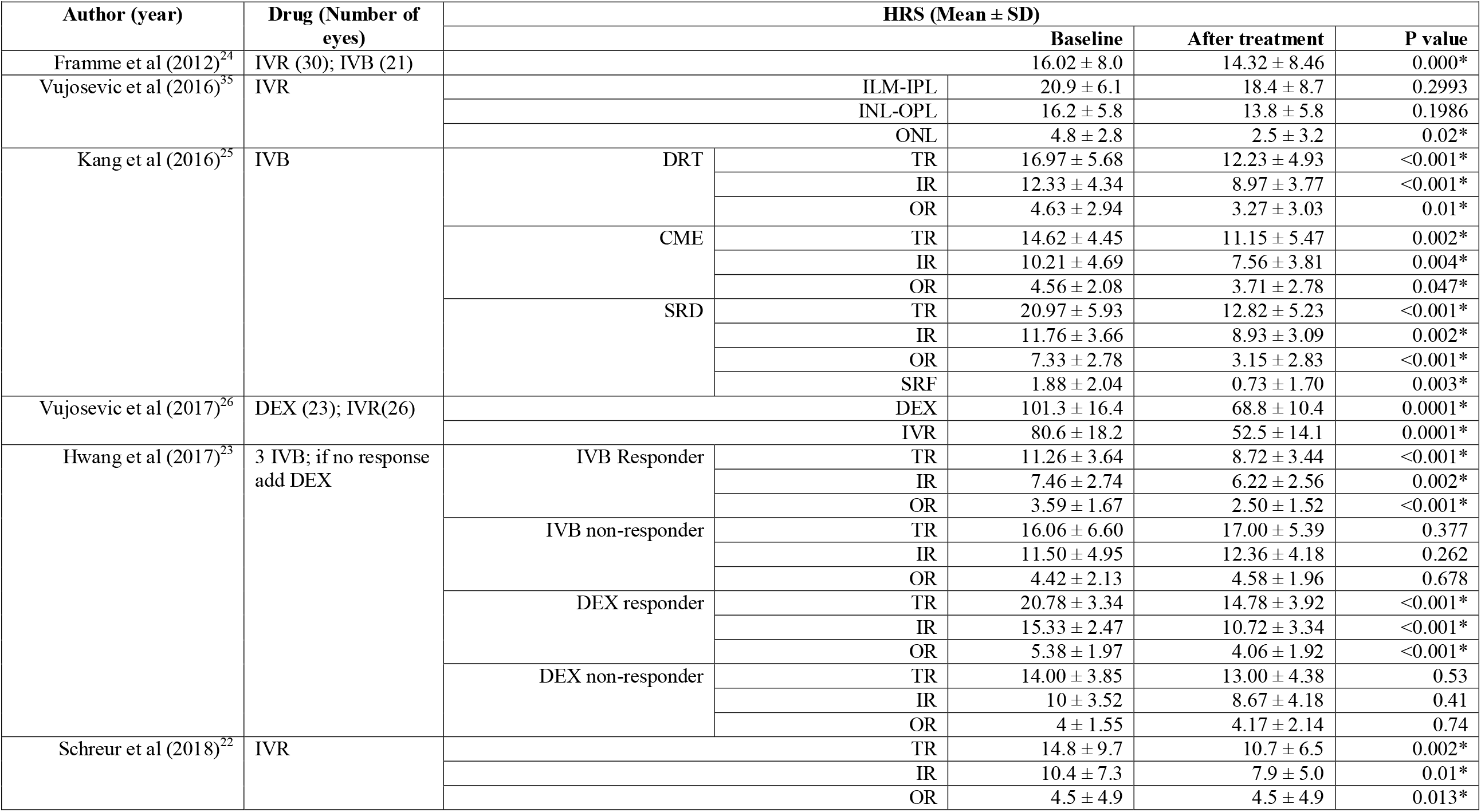
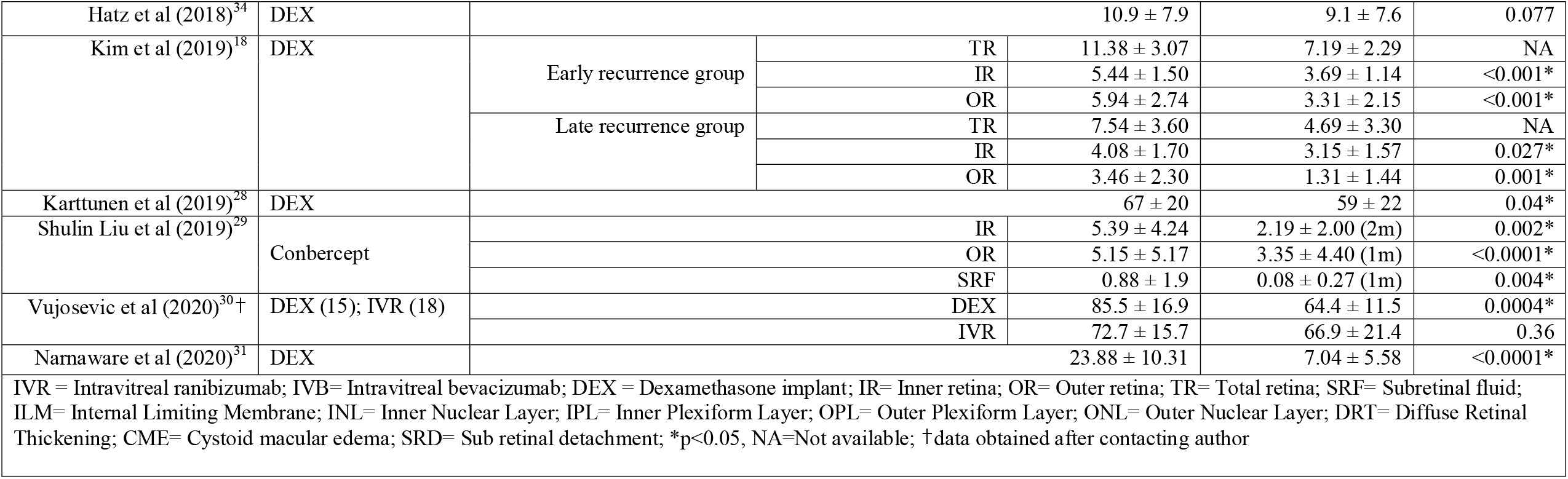
Quantitative HRS Change Following Intravitreal Therapy.

All 7 studies where intravitreal anti-VEGF injections were used^16,17,30-32,35,40^ and 6 out of the 7 studies where dexamethasone was used^17,26,32,34,37,39^ reported a decrease in quantitative HRS. In the subgroup of patients whose macular edema did not respond to dexamethasone or IVB, there was no significant HRS reduction.^17^

Layer-wise analysis of HRS was done in 6 studies. However, the definition of inner and outer retinal layers was variable across the studies. Inner retina (IR) was defined as extending from internal limiting membrane (ILM) to outer nuclear layer (ONL) in 3 studies,^17,31,35^ ILM to inner nuclear layer (INL) in 1 study^26^ and as INL in 1 study.^16^ Similarly, outer retina (OR) was defined as extending from external limiting membrane (ELM) to RPE in 2 studies,^17,31^ ELM to photoreceptors in 1 study^35^ and ELM to outer plexiform layer (OPL) in 2 studies.^16,26^ One study analyzed HRS in three layers i.e., ILM to inner plexiform layer (IPL), INL to OPL and ONL.^40^

### HRS change in steroid versus anti-VEGF treated eyes (Table 2 and Figure 2A)

Two studies compared the change in HRS counts between these two classes of drugs, in treatment naive eyes.^32,36^ Vujosevic et al^32^ showed a greater reduction in HRS in dexamethasone treated eyes (n=15) versus IVR treated eyes (n=18) (24.7% versus 8.0%, p = 0.03) when all baseline parameters were matched. In another study by the same author, the decrease in HRS was not found to be different between the two treatment groups (p=0.135).^36^ However, note that in this study, the baseline HRS counts were significantly higher in the dexamethasone group compared to the IVR group (p=0.003). Hwang et al^17^ noted that baseline HRS numbers were higher in eyes that did not respond to IVB. When such eyes were treated with dexamethasone implant, a significant reduction in HRS was noted.

**Figure 2:**
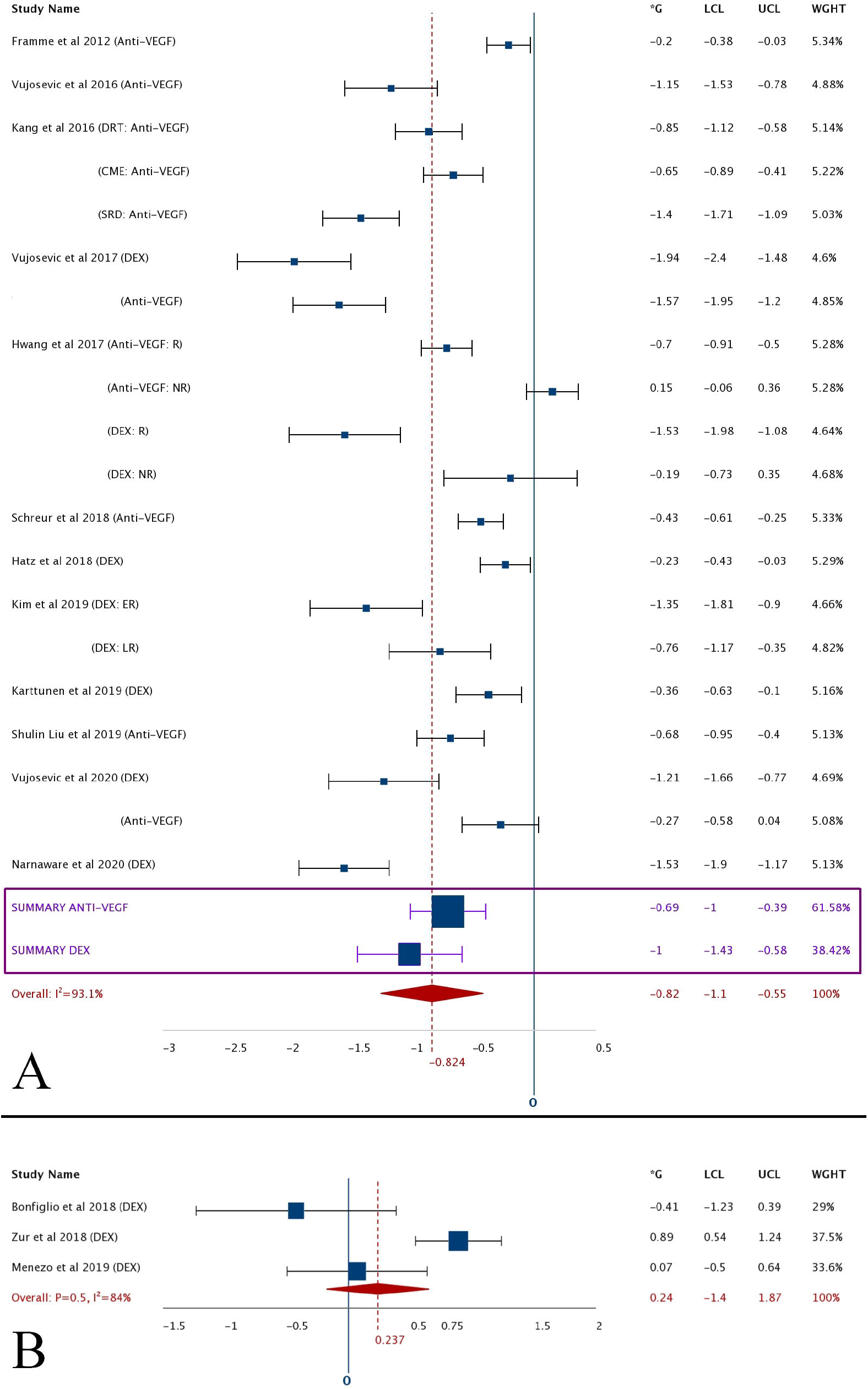
A) Forest plot showing the change in quantitative HRS following intravitreal injection. There were a total of 12 studies among which there were 20 effect sizes to be analyzed. The box and whisker plot for individual studies represent the effect size (Hedges’ g) and 95% confidence intervals (CI_95%_). Subgroup analysis for dexamethasone and anti-VEGF groups are summarized within the plot. The overall effect size is represented by the polygon. B) Forest plot showing the association between HRS at baseline and change in VA. [*G=Hedges’ g; LCL=lower confidence limit; UCL=upper confidence limit; WGHT=weight of the study; dotted vertical line=overall effect size; I^2^=heterogeneity of the studies; within parenthesis=therapeutic group; VEGF=vascular endothelial growth factor; DEX=dexamethasone; DRT=diffuse retinal thickening; CME=cystoid macular edema; SRD=serous retinal detachment; R=responder; NR=non-responder; ER=early recurrence; LR=late recurrence]

### Baseline HRS and Change in VA (Table 3 and Figure 2B)

A total of 14 studies were analyzed. Five studies made a qualitative reporting of HRS as present or absent at baseline^12,14,24,38,41^ while in the rest a quantitative assessment of HRS was done. Of these, 3 studies had categorized the patients into those with HRS <10-15 and those with HRS >10-15 on baseline scans.^15,33,39^ In the rest 6 studies, baseline HRS counts were correlated with final VA using regression/correlation statistics.^16,30,31,35,37,40^

**Table 3:**
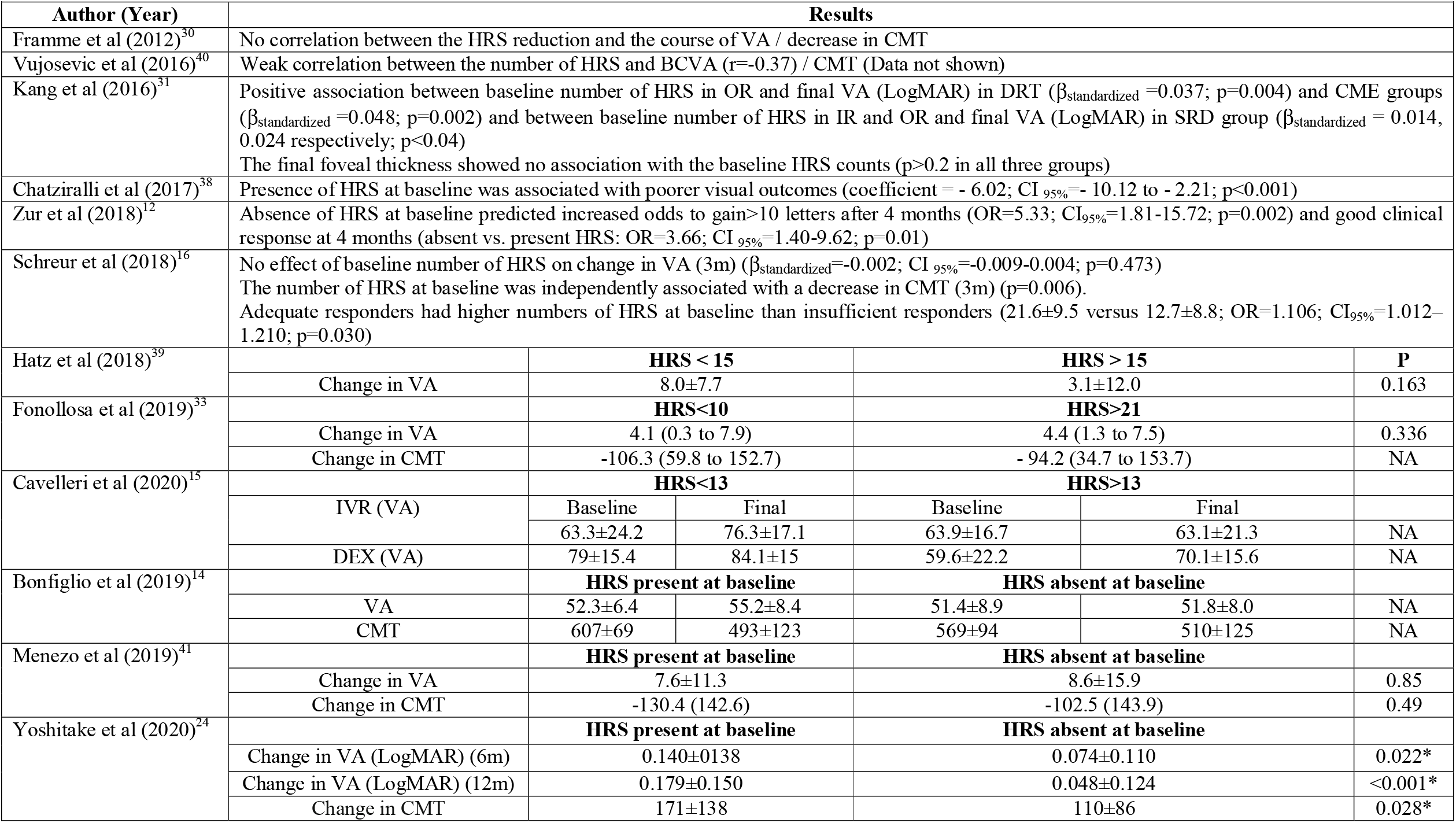
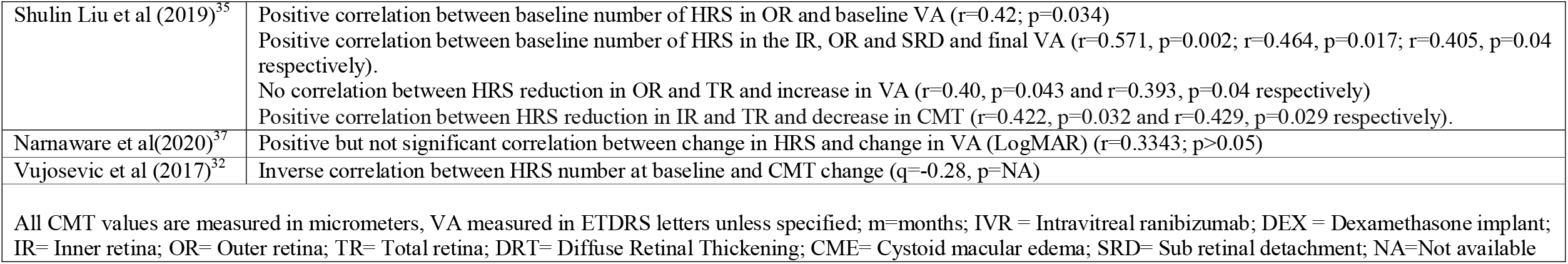
Association between HRS and VA/ CMT.

Three studies showed that higher HRS counts at baseline were associated with worse final VA.^31,35,38^ Five studies showed no correlation between baseline HRS counts and final VA.^15,16,33,39,40^ In a study by Cavalleri et al,^15^ dexamethasone therapy resulted in a greater gain in VA in eyes with high baseline HRS counts compared to IVB. Bonfiglio et al^14^ and Yoshitake et al^24^ compared eyes with and without HRS at baseline and showed a greater gain in VA following dexamethasone and anti-VEGF injections respectively in eyes with HRS. Zur et al^12^ reported a greater gain in eyes without HRS at baseline^12^ and Menezo et al^41^ showed no association between gain in VA and the presence of HRS at baseline.

### Baseline HRS and CMT change (Table 3)

A total of 10 studies were included for this analysis. In 3 studies, the patients were grouped as those with or without HRS at baseline.^14,24,41^ Bonfiglio et al^14^ and Yoshitake et al^24^ reported greater reduction in CMT in eyes with HRS compared to those without. Menezo et al^41^ found no association between the two parameters. Two studies which evaluated the association between a decrease in HRS and change in CMT showed contrasting results, with Liu et al^35^ reporting a significant correlation between the reduction in inner and total retinal HRS and the decrease in CMT at 3 months (r=0.422, p=0.032 and r=0.429, p=0.029 respectively) and Framme et al^30^ reporting no significant association between the two variables at the end of 1 month. Vujosevic et al^32^ showed greater CMT reduction in eyes with more HRS (>87) at baseline than those with less HRS (<87) (ρ=-0.28, p=not reported). Schreur et al^16^ reported that the number of HRS at baseline was independently associated with a decrease in CMT (β_standardized_=-2.61, p=0.006). On the contrary, Fonollosa et al^33^ found that the CMT reduction was not significantly different between groups with scare (<10) or abundant (>21) HRS. Finally, Kang et al^31^ and Vujosevic et al^40^ found no significant correlation between the baseline HRS counts and the final retinal thickness.

### Meta-analysis

Meta-analysis on 12 studies to analyze the quantitative HRS change following intravitreal therapy (figure 2A), showed high heterogeneity in the studies (I^2^ = 93.16%). Funnel plot showed an asymmetry in distribution and significant publication bias (Δ= −100; Kendell’s Tau a= −0.526, CI_95%_= −0.47 to −0.36, PI= −1.57 to 0.73, p=0.001). There was no significant difference between dexamethasone (Hedges’g = −1.0, CI_95%_ = −1.42 to −0.57) and anti-VEGF groups (Hedges’g= −0.69, CI_95%_= −0.99 to −0.38) in terms of HRS reduction (Q*=1.4, df=1, p=0.23).

Due to the heterogeneity in defining the layers, layer-wise changes were not analyzed in meta-analysis. To analyze the association between HRS and VA, we performed a meta-analysis on three studies (figure 2B).^12,14,41^ Presence/absence of HRS at the start of the therapy was not associated with improved VA at the end of treatment (Hedges’ g= 0.237, CI_95%_= −1.39 to 1.87, I^2^=84%, p=0.5) (Figure 2B).

We could not perform a meta-analysis to see the effect of HRS on CMT reduction due to wide heterogeneity in the way the data was reported.

## Discussion

In this review, we found that there is a definite reduction in HRS counts following intravitreal therapy with no significant difference between anti-VEGF and steroid groups. The role of HRS in predicting visual outcome and CMT change was limited by the number of analyzable studies owing to the wide variation in the study designs, methods and reporting.

OCT as an investigating modality has revolutionized the treatment of DME. So far, the response to therapy was being predicted based on the type of edema with centre-involving CME having the worst prognosis owing to the permanent structural damage to the neural elements in the fovea. However, recent research has identified additional biomarkers like NSD, HRS, EZ loss, ELM loss and DRIL to estimate the severity of the edema, decide the type of intravitreal drug to be used or predict functional and anatomical responses to intravitreal therapy.^10-14^

Various theories have been proposed regarding the exact nature of HRS.^18-22^ Of these, the hard exudate and inflammatory theories are most popular in DME. Cusick et al using immunochemistry found apolipoprotein-B deposits corresponding to the HRS in OCT sections.^43^ Intravitreal dexamethasone is a potent anti-inflammatory agent. Anti-VEGF injections, although not as potent as steroids in their anti-inflammatory action, have been shown to have anti-activated microglial activity in addition to antiangiogenic property.^44^ The reduction or disappearance of HRS within 3 months of starting intravitreal therapy as seen in most studies of this review is a strong pointer towards their inflammatory origin. It may be hard to explain such rapid regression of well-formed hard exudates but at this stage the behavioral dynamics of the precursors of hard exudates (HRS) are still unknown.

The results of the meta-analysis did not show a significant effect of baseline HRS on the change in visual acuity. However, eyes with treatment naive DME^15,16,30-32,35,37,40^ as against those with refractory DME^14,38,39^ showed a positive correlation between HRS with VA gain (5/8 studies versus 0/3 studies) and CMT reduction (7/8 studies versus 1/3 studies). Also, eyes where HRS was associated with NSD at baseline responded better and showed a positive correlation between HRS and VA improvement and CMT reduction with in first 2-3 months.^14,31,36,40^ Based on the above observations, we can hypothesize that inflammation plays a dominant role in treatment naive eyes or eyes with NSD when they present with HRS as evidenced by their prompt response to intravitreal injections.

The limitations of the studies included in this review are: retrospective design of most of these studies; small sample sizes in most; lack of uniformity in HRS evaluation as an outcome (variable macular areas over which HRS was analyzed, manual versus automated counting, variability in the definition of IR, OR and TR and masking of investigators); short duration of follow up; lack of adjustment for confounders like [blood lipid and sugar levels,^45,46^ type of macular edema (CME, DRT, SRD)], lack of uniformity in reporting the statistics and outcomes and significant publication bias. Also, there has been a lack of uniform definition of HRS in these studies. Framme et al^21^ described them as “hyperreflective foci of round or oval shape and of different sizes.” Vujosevic et al^40^ described HRS as “small, punctiform discrete white lesions” with no reference to the exact size or reflectivity. Fonollosa et al^33^ cited de Benedetto et al^18^, who stated “small, round- or oval-shaped, well-circumscribed particles (no bigger than 40 μm in diameter), with higher reflectivity than the background.” Yoshitake et al^24^ considered “round or oval particles seen on the OCT images and corresponded to hard exudates in the fundus photographs” as HRS. Cavalleri et al^15^ considered the following morphologic characteristics to define HRS: reflectivity similar to that of nerve fiber layer; absence of back-shadowing; <30 μm diameter. No definition of the HRS were provided by Zur et al^12^, Chatziralli et al^38^, or Kang et al^31^. There is a need to standardize a number of factors before we understand these lesions. Hence, we recommend a stage-wise approach to understand the exact nature and role of this biomarker in DME. (Table 4)

**Table 4:**
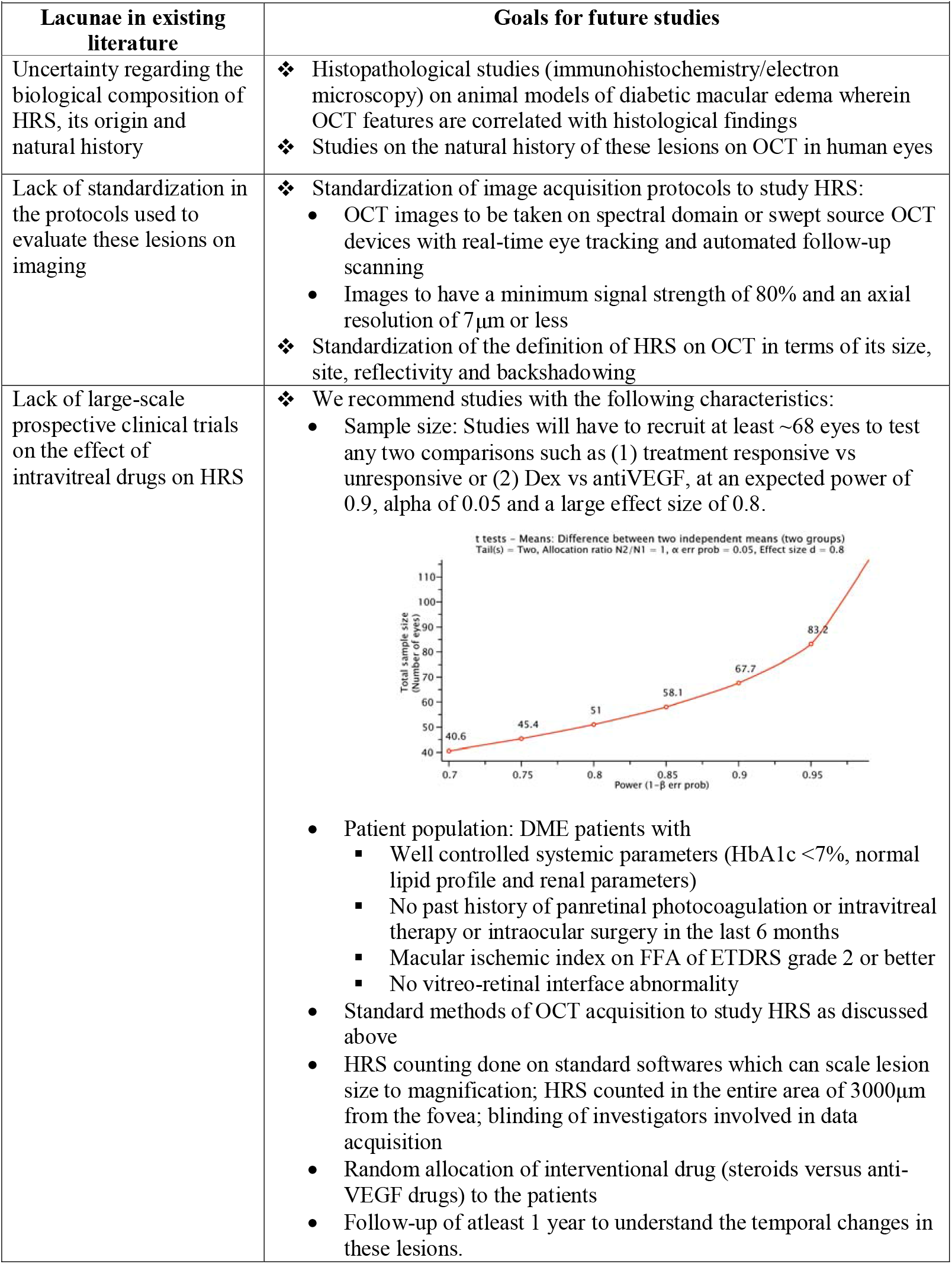
Lacunae in existing literature and goals for future studies.

In conclusion, HRS as a biomarker has been shown to be very promising in predicting therapeutic response to intravitreal treatment in DME in the individual studies. This review could conclude that there is a definite reduction in quantitative HRS following either form of intravitreal therapy. But, owing to the high heterogeneity in reporting, this meta-analysis could not summarize the predictive value of HRS in determining the VA and CMT outcomes.

## Supporting information

Search Strategy

## Data Availability

This is a meta-analysis

## Acknowledgements

We thank Dr. Stela Vujosevic, MD, PhD, University Hospital Maggiore della Carità, Corso Mazzini 18, 28100 Novara, Italy and Dinah Zur, MD, Tel Aviv University, Tel Aviv, Israel for sharing the original data in order to conduct this meta-analysis.

## Appendix 1

Complete search strategy for PubMed (searched on 4th July 2020)

## References

1. Sun JK, Jampol LM. The Diabetic Retinopathy Clinical Research Network (DRCR.net) and Its Contributions to the Treatment of Diabetic Retinopathy. Ophthalmic Res. 2019;62:225–230.

2. Nguyen QD, Brown DM, Marcus DM, et al. Ranibizumab for Diabetic Macular Edema: Results from 2 Phase III Randomized Trials: RISE and RIDE. Ophthalmology. 2012;119:789–801.

3. Gonzalez VH, Campbell J, Holekamp NM, et al. Early and Long-Term Responses to Anti-Vascular Endothelial Growth Factor Therapy in Diabetic Macular Edema: Analysis of Protocol I Data. Am J Ophthalmol. 2016;172:72–79.

4. Bressler NM, Beaulieu WT, Glassman AR, et al. Persistent macular thickening following intravitreous aflibercept, bevacizumab, or ranibizumab for central-involved diabetic macular edema with vision impairment: a secondary analysis of a randomized clinical trial. JAMA Ophthalmol 2018;136:257–269

5. Busch C, Fraser-Bell S, Iglicki M, et al. Real-world outcomes of non-responding diabetic macular edema treated with continued anti-VEGF therapy versus early switch to dexamethasone implant: 2-year results. Acta Diabetol. 2019;56:1341–1350.

6. Regillo CD, Callanan DG, Do DV, et al. Use of Corticosteroids in the Treatment of Patients With Diabetic Macular Edema Who Have a Suboptimal Response to Anti-VEGF: Recommendations of an Expert Panel. Ophthalmic Surg Lasers Imaging Retina. 2017;48:291–301.

7. Funatsu H, Noma H, Mimura T, et al. Association of Vitreous Inflammatory Factors with Diabetic Macular Edema. Ophthalmology. 2009;116:73–9.

8. Sonoda S, Sakamoto T, Yamashita T, et al. Retinal morphologic changes and concentrations of cytokines in eyes with diabetic macular edema. Retina. 2014;34:741–748.

9. Choi MY, Jee D, Kwon JW. Characteristics of diabetic macular edema patients refractory to anti-VEGF treatments and a dexamethasone implant. PLoS ONE. 2019;14:e0222364.

10. Gerendas BS, Prager S, Deak G, et al. Predictive imaging biomarkers relevant for functional and anatomical outcomes during ranibizumab therapy of diabetic macular oedema. Br J Ophthalmol. 2018;102:195–203.

11. Sheu S-J, Lee Y-Y, Horng Y-H, et al. Characteristics of diabetic macular edema on optical coherence tomography may change over time or after treatment. Clin Ophthalmol Auckl NZ. 2018;12:1887–93.

12. Zur D, Iglicki M, Busch C, et al. OCT Biomarkers as Functional Outcome Predictors in Diabetic Macular Edema Treated with Dexamethasone Implant. Ophthalmology. 2018;125:267–275.

13. Sun JK, Lin MM, Lammer J, et al. Disorganization of the Retinal Inner Layers as a Predictor of Visual Acuity in Eyes With Center-Involved Diabetic Macular Edema. JAMA Ophthalmol. 2014;132:1309–1316.

14. Bonfiglio V, Reibaldi M, Pizzo A, et al. Dexamethasone for unresponsive diabetic macular oedema: optical coherence tomography biomarkers. Acta Ophthalmol (Copenh). 2019;97:e540–544.

15. Cavalleri M, Cicinelli MV, Parravano M, et al. Prognostic role of optical coherence tomography after switch to dexamethasone in diabetic macular edema. Acta Diabetol. 2020;57:163–171

16. Schreur V, Altay L, van Asten F, et al. Hyperreflective foci on optical coherence tomography associate with treatment outcome for anti-VEGF in patients with diabetic macular edema. PLOS ONE. 2018;13:e0206482.

17. Hwang HS, Chae JB, Kim JY, Kim DY. Association Between Hyperreflective Dots on Spectral-Domain Optical Coherence Tomography in Macular Edema and Response to Treatment. Invest Ophthalmol Vis Sci. 2017;58:5958–5967.

18. De Benedetto U, Sacconi R, Pierro L, et al. Optical coherence tomographic hyperreflective foci in early stages of diabetic retinopathy. Retina. 2015;35:449–453.

19. Zeng HY, Green WR, Tso MO. Microglial activation in human diabetic retinopathy. Arch Ophthalmol. 2008;126:227–232

20. Altmann C, Schmidt MHH. The Role of Microglia in Diabetic Retinopathy: Inflammation, Microvasculature Defects and Neurodegeneration. Int J Mol Sci. 2018;19:110

21. Framme C, Wolf S, Wolf-Schnurrbusch U. Small dense particles in the retina observable by spectral-domain optical coherence tomography in age-related macular degeneration. Invest Ophthalmol Vis Sci. 2010;51:5965–5969

22. Yoshitake T, Murakami T, Suzuma K, et al. Predictor of Early Remission of Diabetic Macular Edema under As-Needed Intravitreal Ranibizumab. Sci Rep. 2019;9:7599

23. Maggio E, Sartore M, Attanasio M, et al. Anti-Vascular Endothelial Growth Factor Treatment for Diabetic Macular Edema in a Real-World Clinical Setting. Am J Ophthalmol. 2018;195:209–22.

24. Yoshitake T, Murakami T, Suzuma K, et al. Hyperreflective Foci in the Outer Retinal Layers as a Predictor of the Functional Efficacy of Ranibizumab for Diabetic Macular Edema. Sci Rep. 2020;10:873–873.

25. Park YG, Choi MY, Kwon JW. Factors associated with the duration of action of dexamethasone intravitreal implants in diabetic macular edema patients. Sci Rep. 2019;9:19588

26. Kim KT, Kim DY, Chae JB. Association between Hyperreflective Foci on Spectral-Domain Optical Coherence Tomography and Early Recurrence of Diabetic Macular Edema after Intravitreal Dexamethasone Implantation. J Ophthalmol. 2019;2019:3459164

27. Stroup DF, Berlin JA, Morton SC, Olkin I, Williamson GD, Rennie D, Moher D, Becker BJ, Sipe TA, Thacker SB. Meta-analysis of observational studies in epidemiology: a proposal for reporting. Meta-analysis Of Observational Studies in Epidemiology (MOOSE) group. JAMA. 2000 Apr 19;283:2008–12.

28. Study Quality Assessment Tools; National Heart, Lung, and Blood Institute (NHLBI) Available from: https://www.nhlbi.nih.gov/health-topics/study-quality-assessment-tools [accessed on 10th June 2020]

29. Suurmond R, van Rhee H, Hak T. Introduction, comparison, and validation of MetalJEssentials: A free and simple tool for metalJanalysis. Res Synth Methods. 2017;8:537–553.

30. Framme C, Schweizer P, Imesch M, Wolf S, Wolf-Schnurrbusch U. Behavior of SD-OCT-detected hyperreflective foci in the retina of anti-VEGF-treated patients with diabetic macular edema. Invest Ophthalmol Vis Sci. 2012;53:5814–5818

31. Kang JW, Lee H, Chung H, Kim HC. Correlation between optical coherence tomographic hyperreflective foci and visual outcomes after intravitreal bevacizumab for macular edema in branch retinal vein occlusion. Graefes Arch Clin Exp Ophthalmol. 2014;252:1413–21.

32. Vujosevic S, Torresin T, Bini S, et al. Imaging retinal inflammatory biomarkers after intravitreal steroid and anti-VEGF treatment in diabetic macular oedema. Acta Ophthalmol (Copenh). 2017;95:464–471.

33. Fonollosa A, Zarranz-Ventura J, Valverde A, et al. Predictive capacity of baseline hyperreflective dots on the intravitreal dexamethasone implant (Ozurdex®) outcomes in diabetic macular edema: a multicenter study. Graefes Arch Clin Exp Ophthalmol. 2019;257:2381–2390.

34. Karttunen T, Nummelin L, Kaarniranta K, Kinnunen K. Real life experience of dexamethasone implant in refractory diabetic macular oedema. Clin Ophthalmol. 2019;13:2583–90.

35. Liu S, Wang D, Chen F, Zhang X. Hyperreflective foci in OCT image as a biomarker of poor prognosis in diabetic macular edema patients treating with Conbercept in China. BMC Ophthalmol. 2019;19:157.

36. Vujosevic S, Toma C, Villani E, et al. Diabetic macular edema with neuroretinal detachment: OCT and OCT-angiography biomarkers of treatment response to anti-VEGF and steroids. Acta Diabetol. 2020;57:287–296.

37. Narnaware SH, Bawankule PK, Raje D. Short-term outcomes of intravitreal dexamethasone in relation to biomarkers in diabetic macular edema. Eur J Ophthalmol. 2020;1120672120925788.

38. Chatziralli I, Theodossiadis P, Parikakis E, et al. Dexamethasone Intravitreal Implant in Diabetic Macular Edema: Real-Life Data from a Prospective Study and Predictive Factors for Visual Outcome. Diabetes Ther. 2017;8:1393–1404.

39. Hatz K, Ebneter A, Tuerksever C, et al. Repeated Dexamethasone Intravitreal Implant for the Treatment of Diabetic Macular Oedema Unresponsive to Anti-VEGF Therapy: Outcome and Predictive SD-OCT Features. Ophthalmol Int J Ophthalmol. 2018;239:205–214.

40. Vujosevic S, Berton M, Bini S, et al. Hyperreflective retinal spots and visual function after anti-vascular endothelial growth factor treatment in center-involving diabetic macular edema. Retina. 2016;36:1298–1308.

41. Menezo M, Roca M, Menezo V, Pascual I. Intravitreal dexamethasone implant Ozurdex in the treatment of diabetic macular edema in patients not previously treated with any intravitreal drug: a prospective 12-month follow-up study. Curr Med Res Opin. 2019;35:2111–2106.

42. Chatziralli IP, Sergentanis TN, Sivaprasad S. Hyperreflective foci as an independent visual outcome predictor in macular edema due to retinal vascular diseases treated with intravitreal dexamethasone or ranibizumab. Retina. 2016;36:2319–28.

43. Cusick M, Chew EY, Chan C-C, et al. Histopathology and regression of retinal hard exudates in diabetic retinopathy after reduction of elevated serum lipid levels. Ophthalmology. 2003;110:2126–2133.

44. Forstreuter F, Lucius R, Mentlein R. Vascular endothelial growth factor induces chemotaxis and proliferation of microglial cells. J Neuroimmunol. 2002;132:93–8.

45. Wong BS, Sharanjeet-Kaur S, Ngah NF, Sawri RR. The Correlation between Hemoglobin A1c (HbA1c) and Hyperreflective Dots (HRD) in Diabetic Patients. Int J Environ Res Public Health. 2020;17:3154

46. Bolz M, Schmidt-Erfurth U, Deak G, et al. Optical Coherence Tomographic Hyperreflective Foci: A Morphologic Sign of Lipid Extravasation in Diabetic Macular Edema. Ophthalmology. 2009;116:914–20.

