## Supplementary material for "Role of hyper-reflective spots in predicting outcomes of intravitreal therapy in diabetic macular edema: A systematic review and meta-analysis": Search Strategy

| Appendix:1- Pubmed search strategy ( searched on 4^th^ June 2020) | | | |
| --- | --- | --- | --- |
| No. | Search no | Query | Results |
| 1. | #S1 | Search ((((“DMO” OR “DME” OR “macular oedema” OR “Macular edema” OR “Center-involving” OR “Maculopathy”))) OR ((“DMO”[Title/Abstract] OR “DME”[Title/Abstract] OR “macular oedema”[Title/Abstract] OR “Macular edema”[Title/Abstract] OR “Center-involving”[Title/Abstract] OR “Maculopathy”)[Title/Abstract])) OR ((“DMO” OR “DME” OR “macular oedema” OR “Macular edema” OR “Center-involving” OR “Maculopathy”)[MeSH Terms]) Filters: Humans | [14561](https://pmlegacy.ncbi.nlm.nih.gov/pubmed/?cmd=HistorySearch&querykey=9) |
| 2. | #S2 | Search ((((“Bevacizumab” OR “Optical coherence tomography” OR “Anti VEGF” OR “ranibizumab” OR “SD-OCT” OR “Intravitreal” OR “antivascular endothelial growth factor” OR “OCT” OR “BVZ” OR “dexamethasone” OR “steroid” OR “Intra vitreal” OR “Intra-vitreal” OR “avastin” OR “Lucentis” OR “accentrix” OR “Aflibercept” OR “Eyelea” OR “ozurdex” OR “Triamcinolone acetonide” OR “IVTA” OR “Conbercept” OR “Anti-VEGF” OR “AntiVEGF”))) OR ((“Bevacizumab” OR “Optical coherence tomography” OR “Anti VEGF” OR “ranibizumab” OR “SD-OCT” OR “Intravitreal” OR “antivascular endothelial growth factor” OR “OCT” OR “BVZ” OR “dexamethasone” OR “steroid” OR “Intra vitreal” OR “Intra-vitreal” OR “avastin” OR “Lucentis” OR “accentrix” OR “Aflibercept” OR “Eyelea” OR “ozurdex” OR “Triamcinolone acetonide” OR “IVTA” OR “Conbercept” OR “Anti-VEGF” OR “AntiVEGF”)[MeSH Terms])) OR ((“Bevacizumab”[Title/Abstract] OR “Optical coherence tomography”[Title/Abstract] OR “Anti VEGF”[Title/Abstract] OR “ranibizumab”[Title/Abstract] OR “SD-OCT”[Title/Abstract] OR “Intravitreal”[Title/Abstract] OR “antivascular endothelial growth factor”[Title/Abstract] OR “OCT”[Title/Abstract] OR “BVZ”[Title/Abstract] OR “dexamethasone”[Title/Abstract] OR “steroid”[Title/Abstract] OR “Intra vitreal”[Title/Abstract] OR “Intra-vitreal”[Title/Abstract] OR “avastin”[Title/Abstract] OR “Lucentis”[Title/Abstract] OR “accentrix”[Title/Abstract] OR “Aflibercept”[Title/Abstract] OR “Eyelea”[Title/Abstract] OR “ozurdex”[Title/Abstract] OR “Triamcinolone acetonide”[Title/Abstract] OR “IVTA”[Title/Abstract] OR “Conbercept”[Title/Abstract] OR “Anti-VEGF”[Title/Abstract] OR “AntiVEGF”)[Title/Abstract]) Filters: Humans | [1738178](https://pmlegacy.ncbi.nlm.nih.gov/pubmed/?cmd=HistorySearch&querykey=10) |
| 3. | #S3 | Search ((((“Hyper” OR “reflective” OR “foci” OR “central macular thickness” OR “macular volume” OR “CST” OR “CMT” OR “FT” OR “hyperreflective” OR “foveal thickness” OR “spots” OR “HRS” OR “HF” OR “Small” OR “Dense” OR “Best” OR “Corrected” OR “Visual” OR “acuity” OR “BCVA” OR “outcomes” OR “Hyper-reflective” OR “dots” OR “material” OR “points” OR “aggregates” OR “particles” OR “clumps” OR “retinal” OR “HRF” OR “HS” OR “HRD” OR “inflammatory” OR “biomarkers*” OR “Prognostic” OR “markers”))) OR ((“Hyper” [Title/Abstract] OR “reflective”[Title/Abstract] OR “foci”[Title/Abstract] OR “central macular thickness”[Title/Abstract] OR “macular volume”[Title/Abstract] OR “CST”[Title/Abstract] OR “CMT”[Title/Abstract] OR “FT”[Title/Abstract] OR “hyperreflective”[Title/Abstract] OR “foveal thickness”[Title/Abstract] OR “spots”[Title/Abstract] OR “HRS” [Title/Abstract] OR “HF”[Title/Abstract] OR “Small”[Title/Abstract] OR “Dense”[Title/Abstract] OR “Best”[Title/Abstract] OR “Corrected”[Title/Abstract] OR “Visual”[Title/Abstract] OR “acuity”[Title/Abstract] OR “BCVA”[Title/Abstract] OR “outcomes”[Title/Abstract] OR “Hyper-reflective”[Title/Abstract] OR “dots”[Title/Abstract] OR “material”[Title/Abstract] OR “points”[Title/Abstract] OR “aggregates”[Title/Abstract] OR “particles”[Title/Abstract] OR “clumps”[Title/Abstract] OR “retinal”[Title/Abstract] OR “HRF”[Title/Abstract] OR “HS”[Title/Abstract] OR “HRD”[Title/Abstract] OR “inflammatory”[Title/Abstract] OR “biomarkers*”[Title/Abstract] OR “Prognostic”[Title/Abstract] OR “markers”)[Title/Abstract])) OR ((“Hyper” OR “reflective” OR “foci” OR “central macular thickness” OR “macular volume” OR “CST” OR “CMT” OR “FT” OR “hyperreflective” OR “foveal thickness” OR “spots” OR “HRS” OR “HF” OR “Small” OR “Dense” OR “Best” OR “Corrected” OR “Visual” OR “acuity” OR “BCVA” OR “outcomes” OR “Hyper-reflective” OR “dots” OR “material” OR “points” OR “aggregates” OR “particles” OR “clumps” OR “retinal” OR “HRF” OR “HS” OR “HRD” OR “inflammatory” OR “biomarkers*” OR “Prognostic” OR “markers”)[MeSH Terms]) Filters: Humans | [3886156](https://pmlegacy.ncbi.nlm.nih.gov/pubmed/?cmd=HistorySearch&querykey=11) |
| 4 | #S1 AND S2 AND S3 | Search ((((((((“DMO” OR “DME” OR “macular oedema” OR “Macular edema” OR “Center-involving” OR “Maculopathy”))) OR ((“DMO”[Title/Abstract] OR “DME”[Title/Abstract] OR “macular oedema”[Title/Abstract] OR “Macular edema”[Title/Abstract] OR “Center-involving”[Title/Abstract] OR “Maculopathy”)[Title/Abstract])) OR ((“DMO” OR “DME” OR “macular oedema” OR “Macular edema” OR “Center-involving” OR “Maculopathy”)[MeSH Terms])) AND Humans[Mesh])) AND ((((((“Bevacizumab” OR “Optical coherence tomography” OR “Anti VEGF” OR “ranibizumab” OR “SD-OCT” OR “Intravitreal” OR “antivascular endothelial growth factor” OR “OCT” OR “BVZ” OR “dexamethasone” OR “steroid” OR “Intra vitreal” OR “Intra-vitreal” OR “avastin” OR “Lucentis” OR “accentrix” OR “Aflibercept” OR “Eyelea” OR “ozurdex” OR “Triamcinolone acetonide” OR “IVTA” OR “Conbercept” OR “Anti-VEGF” OR “AntiVEGF”))) OR ((“Bevacizumab” OR “Optical coherence tomography” OR “Anti VEGF” OR “ranibizumab” OR “SD-OCT” OR “Intravitreal” OR “antivascular endothelial growth factor” OR “OCT” OR “BVZ” OR “dexamethasone” OR “steroid” OR “Intra vitreal” OR “Intra-vitreal” OR “avastin” OR “Lucentis” OR “accentrix” OR “Aflibercept” OR “Eyelea” OR “ozurdex” OR “Triamcinolone acetonide” OR “IVTA” OR “Conbercept” OR “Anti-VEGF” OR “AntiVEGF”)[MeSH Terms])) OR ((“Bevacizumab”[Title/Abstract] OR “Optical coherence tomography”[Title/Abstract] OR “Anti VEGF”[Title/Abstract] OR “ranibizumab”[Title/Abstract] OR “SD-OCT”[Title/Abstract] OR “Intravitreal”[Title/Abstract] OR “antivascular endothelial growth factor”[Title/Abstract] OR “OCT”[Title/Abstract] OR “BVZ”[Title/Abstract] OR “dexamethasone”[Title/Abstract] OR “steroid”[Title/Abstract] OR “Intra vitreal”[Title/Abstract] OR “Intra-vitreal”[Title/Abstract] OR “avastin”[Title/Abstract] OR “Lucentis”[Title/Abstract] OR “accentrix”[Title/Abstract] OR “Aflibercept”[Title/Abstract] OR “Eyelea”[Title/Abstract] OR “ozurdex”[Title/Abstract] OR “Triamcinolone acetonide”[Title/Abstract] OR “IVTA”[Title/Abstract] OR “Conbercept”[Title/Abstract] OR “Anti-VEGF”[Title/Abstract] OR “AntiVEGF”)[Title/Abstract])) AND Humans[Mesh])) AND ((((((“Hyper” OR “reflective” OR “foci” OR “central macular thickness” OR “macular volume” OR “CST” OR “CMT” OR “FT” OR “hyperreflective” OR “foveal thickness” OR “spots” OR “HRS” OR “HF” OR “Small” OR “Dense” OR “Best” OR “Corrected” OR “Visual” OR “acuity” OR “BCVA” OR “outcomes” OR “Hyper-reflective” OR “dots” OR “material” OR “points” OR “aggregates” OR “particles” OR “clumps” OR “retinal” OR “HRF” OR “HS” OR “HRD” OR “inflammatory” OR “biomarkers*” OR “Prognostic” OR “markers”))) OR ((“Hyper” [Title/Abstract] OR “reflective”[Title/Abstract] OR “foci”[Title/Abstract] OR “central macular thickness”[Title/Abstract] OR “macular volume”[Title/Abstract] OR “CST”[Title/Abstract] OR “CMT”[Title/Abstract] OR “FT”[Title/Abstract] OR “hyperreflective”[Title/Abstract] OR “foveal thickness”[Title/Abstract] OR “spots”[Title/Abstract] OR “HRS” [Title/Abstract] OR “HF”[Title/Abstract] OR “Small”[Title/Abstract] OR “Dense”[Title/Abstract] OR “Best”[Title/Abstract] OR “Corrected”[Title/Abstract] OR “Visual”[Title/Abstract] OR “acuity”[Title/Abstract] OR “BCVA”[Title/Abstract] OR “outcomes”[Title/Abstract] OR “Hyper-reflective”[Title/Abstract] OR “dots”[Title/Abstract] OR “material”[Title/Abstract] OR “points”[Title/Abstract] OR “aggregates”[Title/Abstract] OR “particles”[Title/Abstract] OR “clumps”[Title/Abstract] OR “retinal”[Title/Abstract] OR “HRF”[Title/Abstract] OR “HS”[Title/Abstract] OR “HRD”[Title/Abstract] OR “inflammatory”[Title/Abstract] OR “biomarkers*”[Title/Abstract] OR “Prognostic”[Title/Abstract] OR “markers”)[Title/Abstract])) OR ((“Hyper” OR “reflective” OR “foci” OR “central macular thickness” OR “macular volume” OR “CST” OR “CMT” OR “FT” OR “hyperreflective” OR “foveal thickness” OR “spots” OR “HRS” OR “HF” OR “Small” OR “Dense” OR “Best” OR “Corrected” OR “Visual” OR “acuity” OR “BCVA” OR “outcomes” OR “Hyper-reflective” OR “dots” OR “material” OR “points” OR “aggregates” OR “particles” OR “clumps” OR “retinal” OR “HRF” OR “HS” OR “HRD” OR “inflammatory” OR “biomarkers*” OR “Prognostic” OR “markers”)[MeSH Terms])) AND Humans[Mesh]) Filters: Journal Article; Publication date from 2011/01/01 to 2020/06/01; Humans; English | 524 |
